# Proposing a New Standard for Collateral Status Assessment in Acute Ischemic Stroke using Cerebrovascular Radiomics

**DOI:** 10.1101/2025.08.01.25332616

**Authors:** Dimitrios Rallios, Adam Hilbert, Charles Majoie, Wim Van H. van Zwam, Aad van der Lugt, Martin Bendszus, Susanne Bonekamp, Peter Vajkoczy, Orhun U. Aydin, Dietmar Frey, the MR CLEAN investigators

**Author notes:** Corresponding Author Details: Dimitrios Rallios CLAIM - Charité Lab for AI in Medicine. Joint supervision (Orhun U. Aydin, Dietmar Frey).

## Abstract

**Background and Aims:** One of the major determinants of individual functional outcome following a large vessel occlusion (LVO) stroke is the degree of collateral circulation, mostly evaluated on CT angiography (CTA). Many clinical trials have established its importance in selecting patients for endovascular thrombectomy. However, assessment of collateral circulation in acute LVO using CTA is rater-dependent and prone to diagnostic errors. We developed an automated cerebrovascular radiomics pipeline to establish objective collateral scoring.

**Methods:** We retrospectively analyzed admission single-phase CTAs from 343 patients included in the MR CLEAN trial. The data was split and stratified by the collateral-score label into training and validation (n=274) and internal testing (n=69) sets. Vessel information was derived from two regions of interest used for radiomics feature extraction: 1) whole cerebral arterial tree, and 2) Circle of Willis artery segments. Segmentation models were developed using the nnU-Net framework on annotated CTAs of the cerebral arterial tree (n = 40) and multiclass Circle of Willis segmentations from the TopCoW dataset (n = 125), respectively. A customized feature selection pipeline identified the most predictive features, which were used to train a random forest classifier to predict collateral status (sufficient: >50% vs. insufficient: <50% filling). We compared the arteries-based approach to established atlas-based middle cerebral artery (MCA) masks and further validated it on an external dataset of 140 acute LVO patients.

**Results:** The vessel segmentation models accurately annotated cerebral arteries (95HD: 4.4932, average Dice coefficient: 0.8347) and the circle of Willis segments (95HD: 2.2711, average DICE coefficient: 0.8095). After radiomics selection, the best predictive features were identified for the cerebral vasculature (n = 6), the MCA mask (n = 98), and the combination model (n = 32). The Vessel-tree-based radiomics model outperformed the MCA mask approach on both internal (AUC: 0.8809 vs. 0.8160) and external (AUC: 0.8339 vs. 0.6628) test sets. Incorporating radiomics features from Circle of Willis segments further boosted performance, achieving the highest external test set AUC (0.8677).

**Conclusion:** We present an accurate, fully automated, and generalizable cerebrovascular radiomics approach for assessing collateral status from admission computed tomography angiography, supporting time-critical decision-making in acute large vessel occlusion.

## Introduction

Acute large vessel occlusion (LVO) is a severe subtype of ischemic stroke associated with poor clinical outcomes ^1^. Beyond rapid diagnosis, comprehensive clinical and radiographic assessments are essential to guide individualized treatment decisions ^2^. One key imaging parameter is the evaluation of collateral circulation - the brain’s ability to compensate for arterial obstruction by redirecting blood flow through alternative vascular pathways ^3^. Despite an occlusion, patients with robust collateral circulation can maintain cerebral perfusion, which helps limit infarct size and improves overall outcomes ^4^. As such, collateral status has emerged as a crucial prognostic and therapeutic factor, with recent studies highlighting its importance in selecting patients for endovascular thrombectomy ^5,6^.

Given the clinical importance of collateral circulation in acute stroke management, reliable assessment at presentation is essential. CT angiography (CTA), the primary imaging modality for evaluating cerebral vessels in the acute setting, is routinely used to assess collateral status ^7,8^.However, despite efforts toward automation and quantification ^9^, the assessment remains subjective and variable even when performed by experienced reviewers ^10,11^.

To facilitate a more accurate vascular assessment, vessel segmentation tools can be utilized to extract detailed representations of the cerebral vasculature ^12^. Automated segmentation models have demonstrated their value across various pathologies, including ischemic stroke ^13^, offering a more objective and reproducible way to visualize and quantify vascular structures ^14^. However, vessel segmentation models have not been studied extensively concerning their clinical utility in downstream tasks and diagnostic value. Radiomics provides a framework for utilizing vessel segmentations as regions of interest to extract quantitative features, enabling consistent, reproducible, and observer-independent assessment ^15^. This technique has shown promise in stroke diagnosis and outcome prediction by analyzing image-derived patterns and textures ^16^. Beyond statistical analysis, radiomics can be integrated into machine learning models to enhance classification performance in clinical tasks^17^.

In this study, we present a comprehensive pipeline for the automated and accurate prediction of collateral status in CT angiography. We begin by employing automated vessel segmentation tools to delineate vascular structures in CTAs, followed by the extraction of vascular radiomics from these segmentations. We apply a customized radiomics selection pipeline to identify the most predictive features from different subsets of radiomics, which are then used to train a Random Forest classifier for collateral classification. To evaluate the effectiveness of using vessel-specific regions of interest (ROIs), we compare our approach with the established method of utilizing Middle Cerebral Artery (MCA) territory masks, as described in reference ^18^, on both internal and external test sets. We also explore the impact of incorporating radiomics from specific Circle of Willis artery segments.

## Methods

For our study, we analyzed the MR CLEAN Trial dataset retrospectively (n = 502), a randomized, multicenter clinical trial conducted between December 2010 and March 2014. This dataset includes clinical and imaging data from patients with proximal intracranial arterial occlusion of the anterior circulation and was designed to compare intra-arterial treatment (thrombolysis, mechanical treatment, or both) plus standard intravenous alteplase with intravenous treatment alone ^8^. Our selection process for the inclusion of patients adhered to the following criteria:

We included patients with acute proximal anterior circulation large vessel occlusions (LVO) who had CTA scans that captured the entire brain or, at a minimum, scans where the lateral ventricles were visible as a criterion for sufficient presentation of the distal brain circulation. As the characteristics of the CTA imaging for the dataset varied for voxel size and slice thickness, patients with slice thickness below 2.0 mm were selected. DICOMs were then converted into NIfTI format for further processing. (n=343)

This dataset was used for developing the binary cerebral artery tree segmentation model and for the training of the downstream radiomics feature based classifier model.

As an external test set, we included patients from the Stroke Unit at the University of Heidelberg who presented with large vessel occlusion (LVO) between 2018 and 2021 and had admission CTA scans that met our predefined imaging inclusion criteria. Due to the limited number of patients with Tan scores of 0 and 1 in the dataset (n = 41), all available cases within these categories were included. To ensure a representation similar to the ratio of the MR CLEAN data, additional patients with Tan scores of 2 and 3 were subsequently selected at random.

For the Circle of Willis segmentation model, we utilized computed tomography angiography (CTA) scans from the 2024 TopCoW (Topology-Aware Anatomical Segmentation of the Circle of Willis) Challenge dataset (n = 125). This challenge aims to support the development of automated segmentation algorithms for clinical applications. The dataset comprises imaging data from patients admitted to the Stroke Center at the University Hospital Zurich (USZ) in 2018 and 2019 with stroke-related neurological conditions. Manual annotations have been provided for the Circle of Willis segments after manual annotation and evaluation from the challenges expert team of neuroradiologists, neurologists and neurosurgeons. All included scans met our predefined voxel size and image quality criteria. Additional details on patient demographics and scanner parameters are available in the official Challenge publication.

### Image preprocessing

#### Our preprocessing Cropping

To capture only brain slices from the original CTAs, we utilized the mri_synthstrip tool from FreeSurfer to generate a brain mask ^19^. SynthStrip was trained using synthetic data and generalizes to any imaging modality. The brain mask was then applied to crop the original CTA images, ensuring that only relevant slices superior to the foramen magnum were included. (Figure 2. A-B-C)

**Figure 1.**
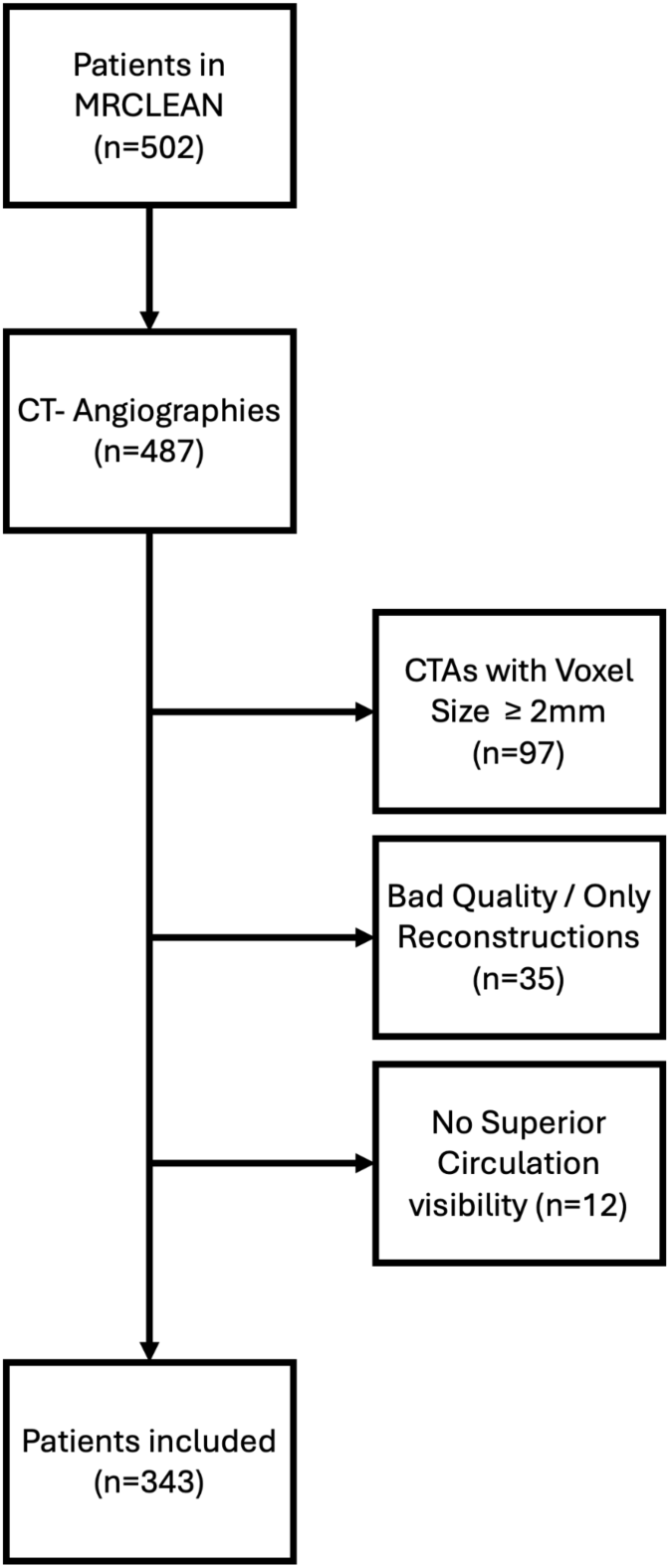
Flow diagram of patient inclusion - MR CLEAN.

**Figure 2.**
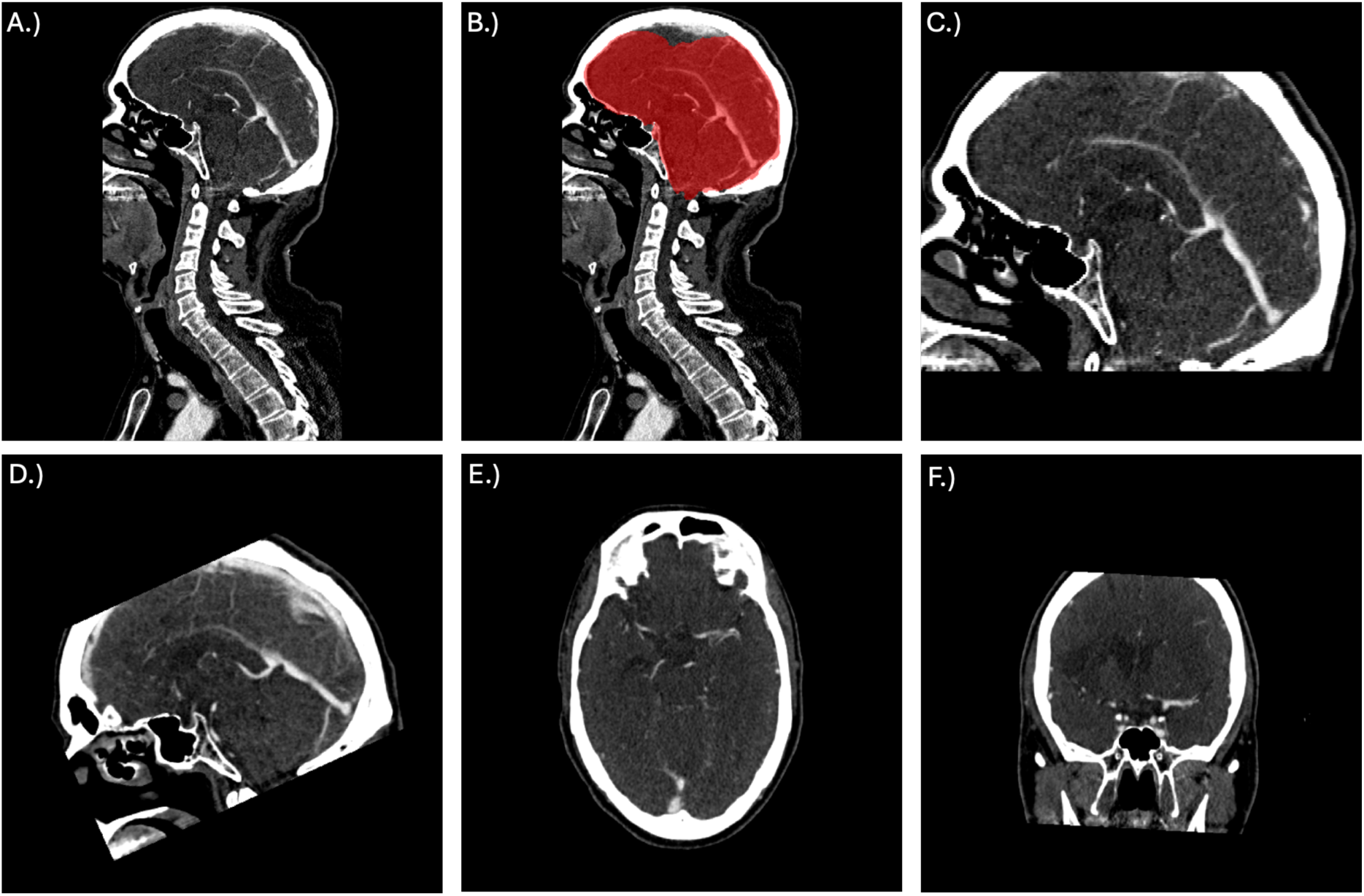
A.) Original image. B.) Generation of the brain mask. C.) Cropping of the initial image guided by the brain mask. D.) Image after template registration (sagittal view). E. Image after template registration, (axial view). F.) Image after template registration (coronal view)

#### Registration

To account for the variation in slice thickness and head orientation between the MR CLEAN and Top CoW scans, we registered CTA images and the vessel segmentation masks from our binary segmentation model to a standardized CT template ^20^. The registration process utilized the FLIRT tool from FSL, which applied a rigid transformation matrix to align voxel sizes and affine matrices with the MNI template. This template had a resolution of 512 x 512 x 512 voxels, each with an isotropic voxel spacing of 0.5 mm, ensuring precise alignment across all data. (Figure 2. D-E-F)

### Vessel Segmentation

We developed two nn-Unet-based ^21^ vessel segmentation models: Model 1, a binary segmentation model for the whole cerebral artery tree, and Model 2, a multiclass model for the Circle of Willis artery segments. The nn-Unet models were trained for 1,000 epochs using the default hyperparameters, with CT normalization and a five-fold cross-validation.

The binary segmentation model (Model 1) was trained through a three-step iterative process. Initially, manual arterial segmentations were performed on CTA images from 10 patients using ITK-SNAP by a junior rater: DR, medical doctor with 2 years of experience in vessel segmentation. The segmentations were then reviewed and refined by two senior raters: OUA, medical doctor with 7 years of experience in vessel segmentation, and DF, neurosurgeon with more than 18 years of clinical experience. This updated dataset of 20 patients was used to train a second nn-UNet model, which was then used to annotate another 20 patients. A second round of expert review and corrections followed this. The process resulted in a dataset of 40 semi-automated segmentations. The third and final nnU-Net model was trained on this dataset of 40 semi-automated segmentations and applied to the entire dataset, achieving a segmentation performance with an average 95th percentile Hausdorff Distance (95HD) of 4.4932 and an average Dice coefficient (DICE) of 0.8347. The whole vessel segmentation masks were split in half, and different labels were attributed to the left and right hemispheres.

For the Circle of Willis segmentation model, we incorporated external annotations from the Top CoW Challenge 2024. The training dataset included 125 CTA images with expert-annotated segmentations of the Circle of Willis, labelled for each Circle of Willis segment: the anterior cerebral artery (ACA), including the A3 segment; the posterior cerebral artery (PCA); the internal carotid artery (ICA); and the middle cerebral artery (MCA). We processed these segmentations to retain distinct labels for the MCA and ICA while consolidating the remaining arteries (ACA, ACoA, PCA, PCoA, and Basilar artery) into a single label. This resulted in a dataset with five distinct vessel labels, which was used to train a separate nnU-Net model. The model achieved an average 95th percentile Hausdorff Distance of 2.2711 and an average Dice coefficient of 0.8095. This model was then applied to predict multi-label segmentations for all patients in the dataset.

We compared our vessel-based study against MCA arterial territory masks derived from vascular lesion distribution. These masks, which have been widely used in previous studies for stroke-related classification tasks ^18^, are part of a comprehensive atlas based on lesion patterns of all cerebral arteries. We modified the original atlas by excluding areas attributed to other arteries and fusing all areas attributed to the MCA and its branches. A detailed map of each segmentation type and modification is shown in Figure 3.

**Figure 3.**
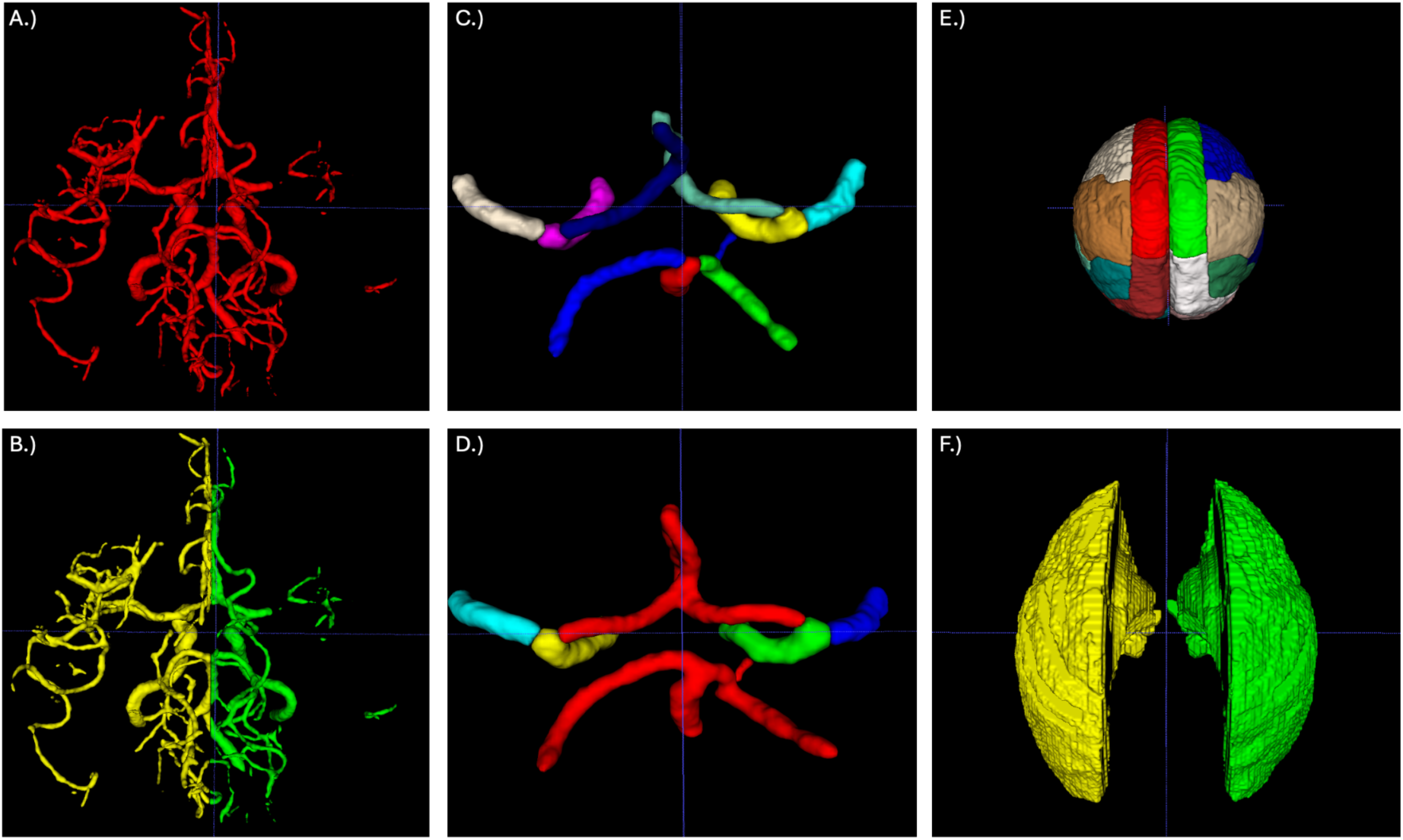
A.) Binary segmentation of the whole cerebral vasculature. B.) Two-labeled segmentation divided for right and left side. C.) Circle of Willis segmentation with individual labelling for each branch. D.) Modified CoW segmentation. E.) Brain atlas showing color-coded labels for regions supplied by each branch of the major arteries F.) Modified MCA Atlas

### Label selection and their classification

In both MR CLEAN trial and the Heidelberg data set, collateral status was assessed using the Tan Collateral scores ^22^, a 4-point grading system that evaluates pial arterial filling distal to an occlusion: grade 0 indicates no collateral filling, grade 1 indicates less than 50% perfusion, grade 2 indicates 50– 99% perfusion, and grade 3 indicates complete or near-complete filling (100%). For model development, collateral status was dichotomized into sufficient (Tan score 2 or 3, n = 274) and non-sufficient (Tan score 0 or 1, n= 69).

For the MR CLEAN trial dataset, collateral grading was performed by two independent and experienced neuroradiologists from the MR CLEAN imaging committee. In cases of disagreement, a third expert reader provided the final decision. All raters were blinded to clinical data, except for the symptomatic side ^23^. In the Heidelberg dataset, collateral status was assessed by a single neuroradiologist from the University Hospital Heidelberg (UKHD).

### Radiomics

In this study, we used the PyRadiomics library ^24^ to extract shape and texture features from CTA images of patients, using in total nine regions of interest: the MCA territory masks (2 - right and left) and cerebral vasculature segmentations (side specific vascular trees (2), right and left MCA (2), right and left MCA (2) and communicating branches (1)). A structured strategy was defined for the extraction process and documented in yaml file format (Appendix 1).

Radiomic features capture the complex patterns of Hounsfield Unit (HU) intensities, which can vary significantly across CT scanners and contrast agent dynamics in CTA. To enhance robustness and ensure generalizability, we standardized all CTA images by clipping voxel intensities to the 1–500 HU range and applying z-score normalization based on each image’s mean and standard deviation. Next, we applied high- and low-pass filters in all spatial directions using the “coif-1” wavelet transform and the Laplacian of Gaussian (LoG) filter for edge enhancement, with sigma values of 2, 4, and 6 mm. Using this configuration, 1,094 radiomic features—including first-order statistics and texture-matrix features — were extracted from each region of interest. For vessel segmentation masks, we additionally extracted shape features (14 shape radiomics).

For the vessel segmentation-based ROIs, we conducted an exploratory analysis of the internal dataset to identify the most predictive subset of features. Specifically, we studied three different subsets of radiomics: 1) only texture features, 2) only shape features, and 3) a combination of both extracted from vessel segmentation masks to identify the best combination (Model A). For the MCA territory-based ROI selection, all radiomic features extracted from the right and left MCA territories were input into the radiomics selection pipeline, resulting in model B. Finally, we combined the best-performing subset of radiomics with Circle of Willis radiomics to produce Model C. The structure of our experimental study is shown in Figure 4.

**Figure 4.**
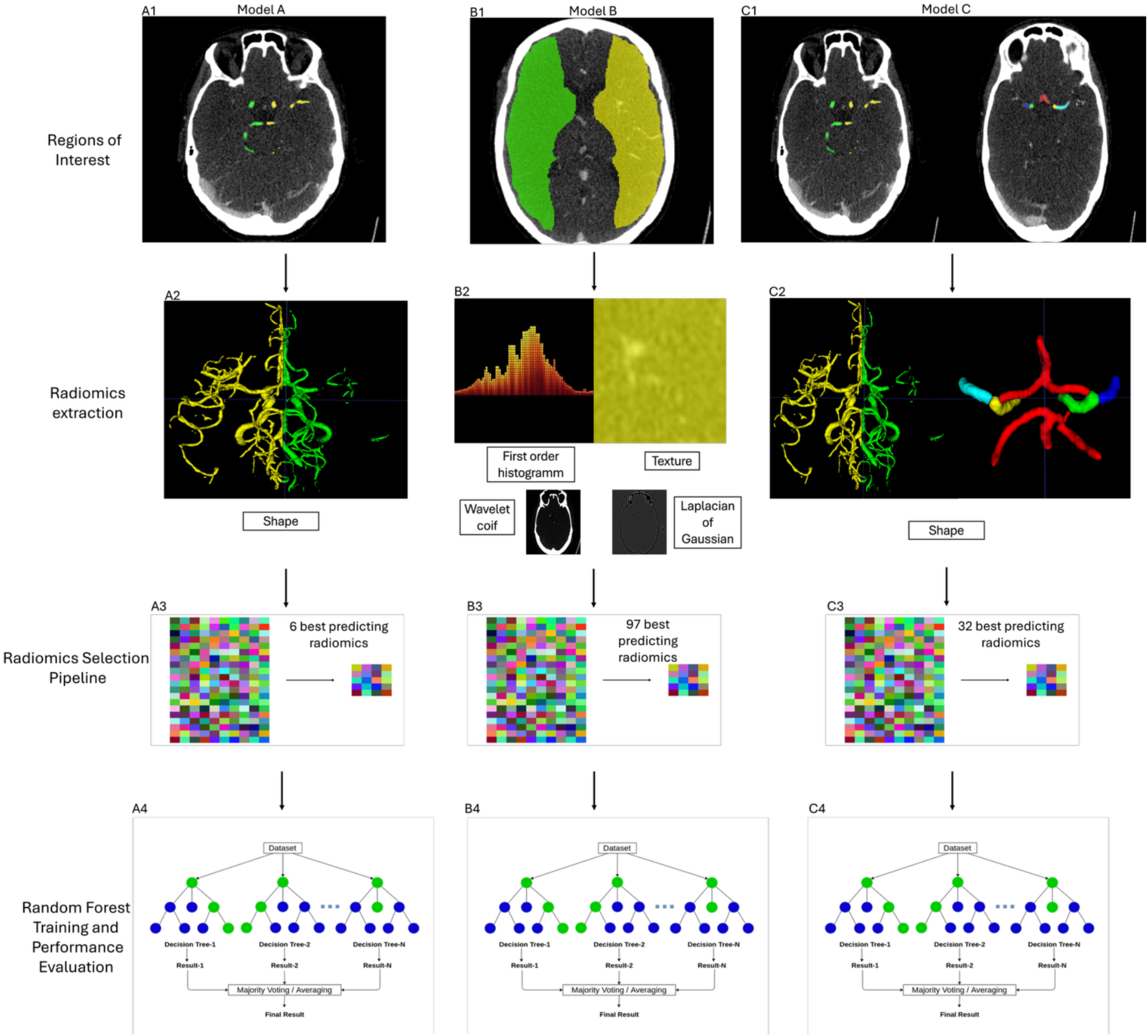
Experimental structure A.) A1.) Patient-specific vessel segmentations are created using the segmentation model, divided into left and right hemispheres, and registered with the original image. A2.) Shape radiomics features are extracted. A3.) The radiomics selection pipeline identifies the best predictive features. A4.) Based on these features, the Random Forest Classifier (RFC) is trained. **B.)** MCA Masks study B1.) MCA masks are registered onto the CTA image. B2.) To enhance texture components, Laplacian of Gaussian (LoG) and wavelet transforms are applied to the CTA. Radiomics features are extracted from both the original and transformed images. B3.) All extracted features pass through a selection pipeline, resulting in a set of the most predictive and non-redundant features. B4.) These selected features are used to train a Random Forest model, with performance evaluated on both internal and external test sets. **C.)** Model C C1.) Patient-specific vessel segmentations (including the specific study on Circle of Willis) are created using the segmentation models. C2.) Shape radiomics features for 7 different ROI (left hemisphere, right hemisphere, left and right ICA, left and right MCA and (the communicating arteries including PCA and ACA were extracted .C3.) The radiomics selection pipeline identifies the best predictive features. 4.) Based on these features, the Random Forest Classifier (RFC) is trained.

Following radiomics feature extraction, we applied min-max normalization to scale all features to a range of 0 to 1. To prevent data leakage, we first split the dataset into training and test sets. Normalization parameters (i.e., the minimum and maximum values for each feature) were computed exclusively from the training set. These values were then saved and used to normalize both internal and external test sets.

### Machine Learning protocol

The machine learning protocol of this study was designed to comply with the CLAIM ^25^ checklist for AI in medical imaging and the CheckList for EvaluAtion of Radiomics research (CLEAR) ^26^.

### Feature selection process

In our study, up to 1,118 radiomic features were extracted for each segmented ROI. Since the selection was based solely on the training set (n = 274), including all radiomics features for classification would create a high-dimensional feature space, potentially increasing the risk of overfitting and compromising model performance ^27^.

To address this challenge of dimensionality, we first removed highly correlated features while retaining the most predictive ones. Feature importance was initially assessed based on the absolute value of coefficients from a task-specific Ridge-regularized logistic regression model with L2 regularization using 10-fold stratified cross-validation and optimized for AUC. For each pair of features with a correlation above 0.9, the feature with the lower importance was discarded.

Using the same approach of ridge regression, we determined the optimal number of features for the classification task. This involved iteratively evaluating model performance using varying subsets of the top-ranked features. Starting with the top 5 most predictive radiomics features, we performed stratified 10-fold cross-validation and computed the average ROC-AUC (Area Under the Receiver Operating Characteristic Curve) across the folds. The subset of radiomics with the best average validation ROC-AUC for the dataset was selected as the most predictive.

Consecutively, we trained a Random Forest classifier ^28,29^ to distinguish between efficient and inefficient collateral status, using the subset of the most predictive radiomics features. The model employs Bayesian optimization with Optuna ^30^ for hyperparameter tuning. Model performance was evaluated using the average ROC-AUC. After 50 optimization trials, the best hyperparameters were selected, and a final Random Forest model with 1000 trees was trained and validated using 4-fold cross-validation. (Figure 5.) For details on the preprocessing, radiomics selection process and Random Forest classifier please refer to the following open-source code repository: https://github.com/claim-berlin/CollateralScore

**Figure 5.**
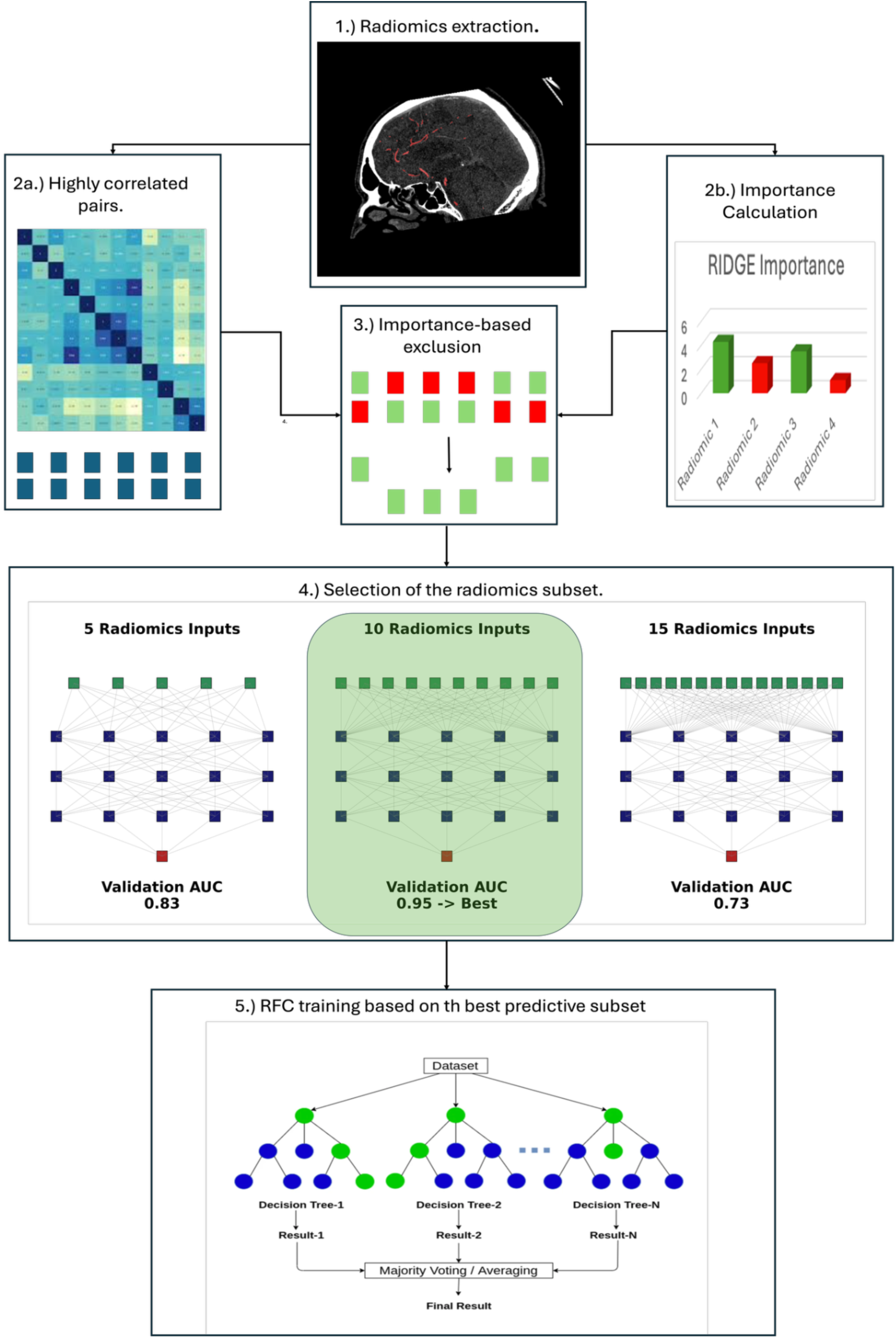
Radiomics selection pipeline: 1.) Extraction of radiomics based on the ROI. 2a.) Calculate correlation between the radiomics and create highly correlated pairs (r>0.9). 2b.) Use a stratified 10-fold RIDGE to calculate feature importance based on the respective task. 3.) Use the importance coefficient to exclude from the pairs the less predictive radiomic.4.) The top predictive Radiomics are incrementally added to optimize Validation AUC. 5.) Random Forest Training based on Radiomics with the best Validation AUC

**Figure 6:**
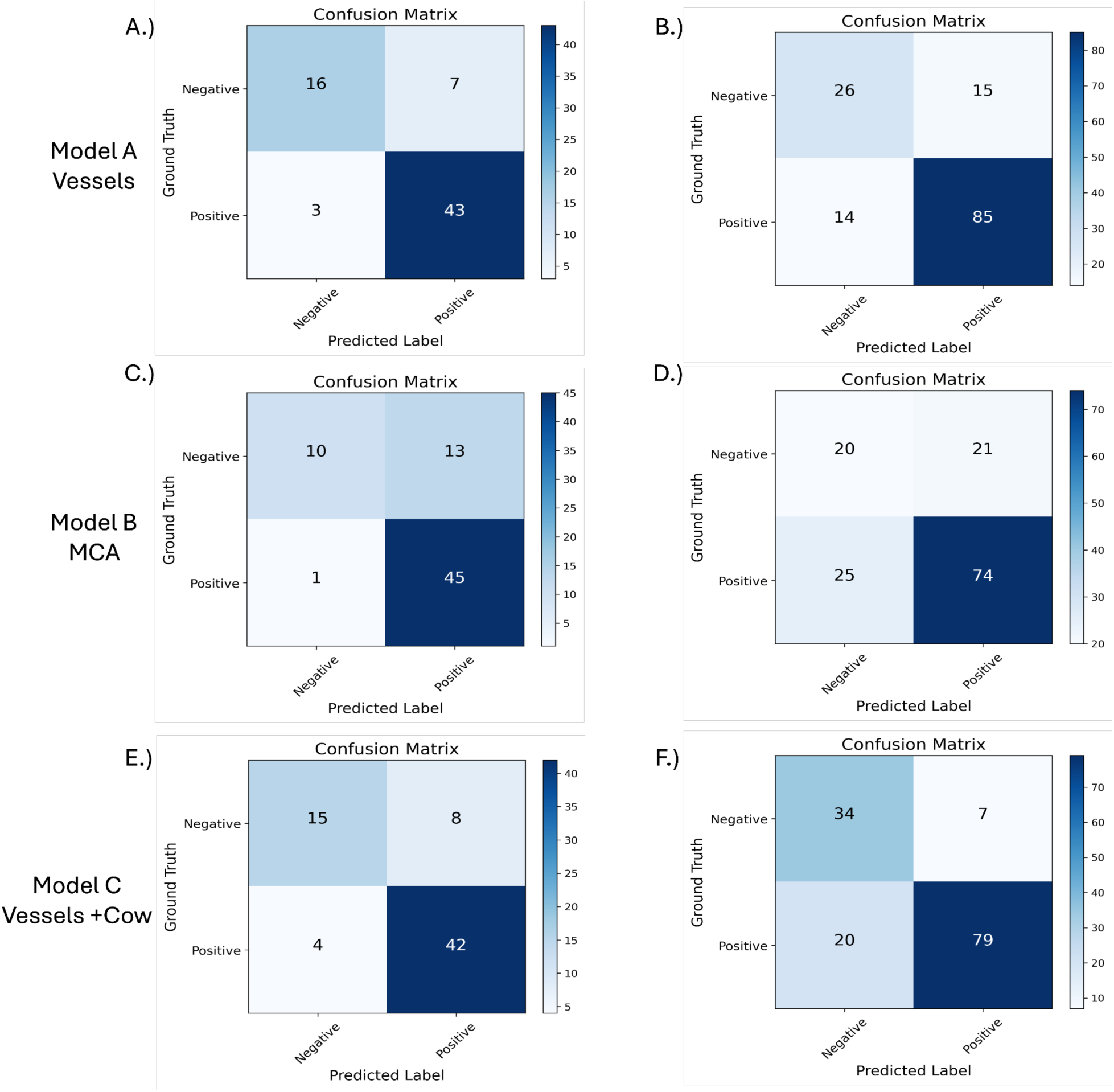
A.) Confusion Matrix Model A - Vessels - Internal Test Set, B.) Confusion Matrix Model A - Vessels - External Test Set, C.) Confusion Matrix Model B - MCA - Internal Test Set D.) Confusion Matrix Model B - MCA - External Test Set D.) Confusion Matrix Model C Vessels + CoW - Internal Test Set, E.) Confusion Matrix Model C f Vessels + CoW - External Test Set

### Statistical analysis and performance metrics

To compare individual clinical variables between the training/cross-validation and independent test sets, we performed univariate statistical analyses. Continuous variables such as age, NIHSS score, and onset-to-CTA time were compared using the student’s *t*-test. Categorical variables, including sex, the hemisphere of symptoms, occlusion site (e.g., M1/M2 vs. ICA), and Collateral score, were evaluated using the Chi-Square exact test. The Area Under the Receiver Operating Characteristic Curve (AUC) was used as the primary performance metric for selecting the most predictive model. To assess the statistical significance of performance differences between models, DeLong’s test was applied. P-values less than 0.05 were considered statistically significant.

## Results

### Patient characteristics

A total of 343 patients were included in the study, with 274 in the training set and 69 in the independent test set. Baseline characteristics, including age, sex, NIHSS score, symptom side, and occlusion site distribution, were well-balanced between groups (all p > 0.05). The time from symptom onset to CTA was slightly shorter in the test set but did not reach statistical significance (p = 0.0915). Detailed patient characteristics are provided in Table 1.A comparison of the internal training set and external test set are provided in Table 2.

**Table 1.**
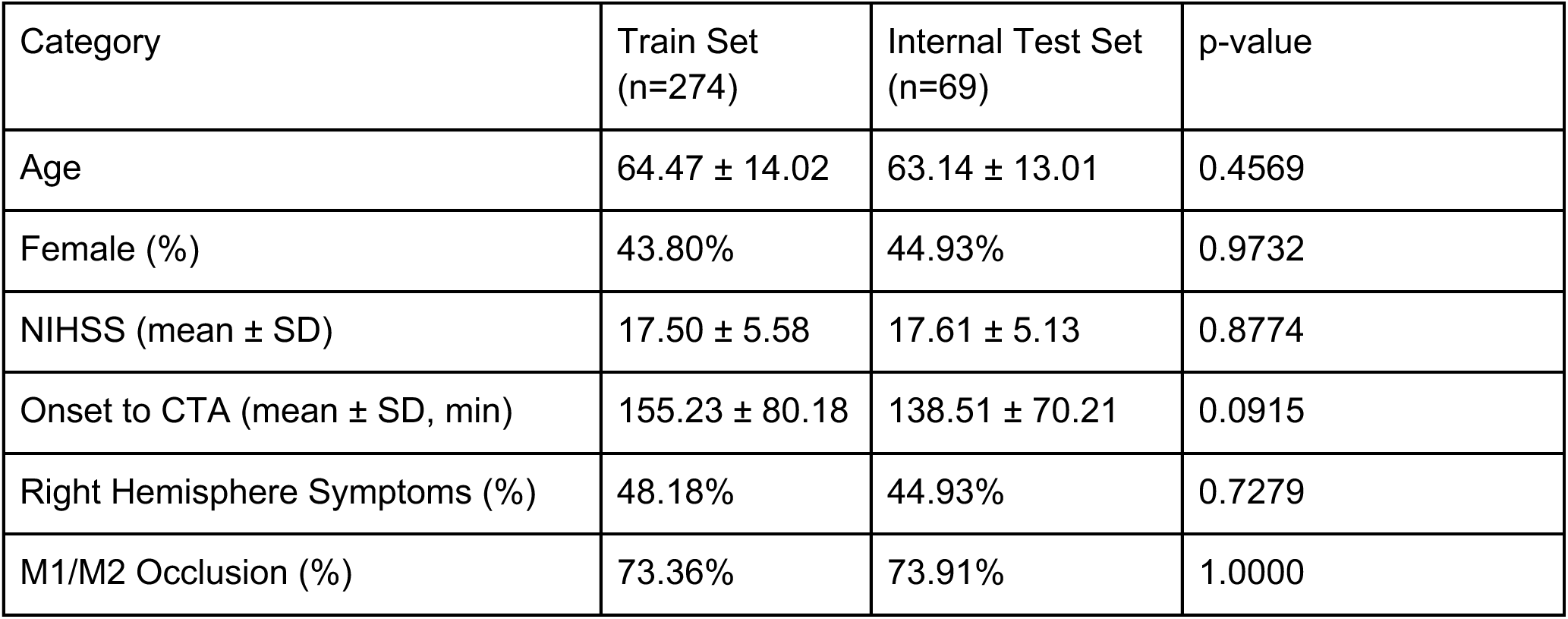
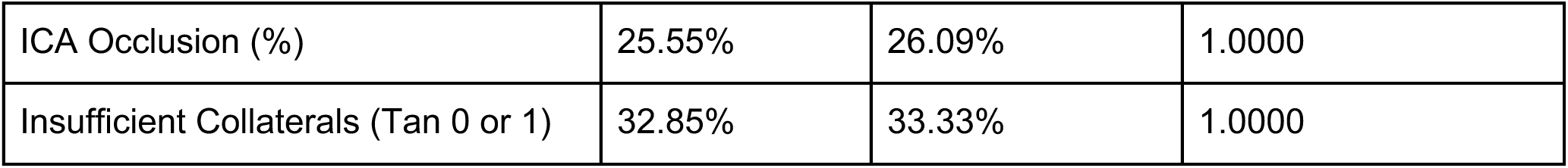
Patient characteristics of Internal Training Set and Internal Test Set.

| Category | Train Set<br>(n=274) | Internal Test Set<br>(n=69) | p-value |
| --- | --- | --- | --- |
| Age | 64.47 ± 14.02 | 63.14 ± 13.01 | 0.4569 |
| Female (%) | 43.80% | 44.93% | 0.9732 |
| NIHSS (mean ± SD) | 17.50 ± 5.58 | 17.61 ± 5.13 | 0.8774 |
| Onset to CTA (mean ± SD, min) | 155.23 ± 80.18 | 138.51 ± 70.21 | 0.0915 |
| Right Hemisphere Symptoms (%) | 48.18% | 44.93% | 0.7279 |
| M1/M2 Occlusion (%) | 73.36% | 73.91% | 1.0000 |
| ICA Occlusion (%) | 25.55% | 26.09% | 1.0000 |
| Insufficient Collaterals (Tan 0 or 1) | 32.85% | 33.33% | 1.0000 |

**Table 2.** Patient characteristics of Internal Training Set and External Test Set.

| Category | Train Set (n=274) | External Test Set (n=140) | p-value |
| --- | --- | --- | --- |
| Age (mean ± SD) | 64.47 ± 14.02 | 72.89 ± 15.33 | <0.0001 |
| NIHSS (mean ± SD) | 17.50 ± 5.58 | 15.27 ± 7.47 | 0.0023 |
| Right Hemisphere Symptoms (%) | 48.18% | 50.71% | 0.7002 |
| M1/M2 Occlusion (%) | 73.36% | 95.00% | <0.0001 |
| ICA Occlusion (%) | 25.55% | 7.14% | <0.0001 |
| Insufficient Collaterals (Tan 0 or 1) | 32.85% | 29.29% | 0.5317 |

#### Binary vessels segmentation ROI

In order to identify the best input for the radiomics selection pipeline, we conducted three initial experiments in the internal test set. First, we used only texture features identical to those from the MCA masks study. The model achieved a test AUC of 0.8440, based on 89 selected features (95% CI: [0.7364, 0.9295]). Second, we fused shape features with the texture features from the first experiment. This pipeline selected 74 features and achieved a test AUC of 0.7779 (95% CI: [0.6571, 0.8888]) on the internal test set. Third, utilizing only shape features (see Figure 7) yielded six selected features. On the internal test set, model A achieved an AUC of 0.8809 (95% CI: [0.7704, 0.9750]), with 43 true positives, 16 true negatives, three false negatives, and seven false positives. In the external test set, the model achieved an AUC of 0.8339 (95% CI: [0.707, 0.909]), with 85 true positives, 26 true negatives, 15 false positives, and 14 false negatives.

**Figure 7:**
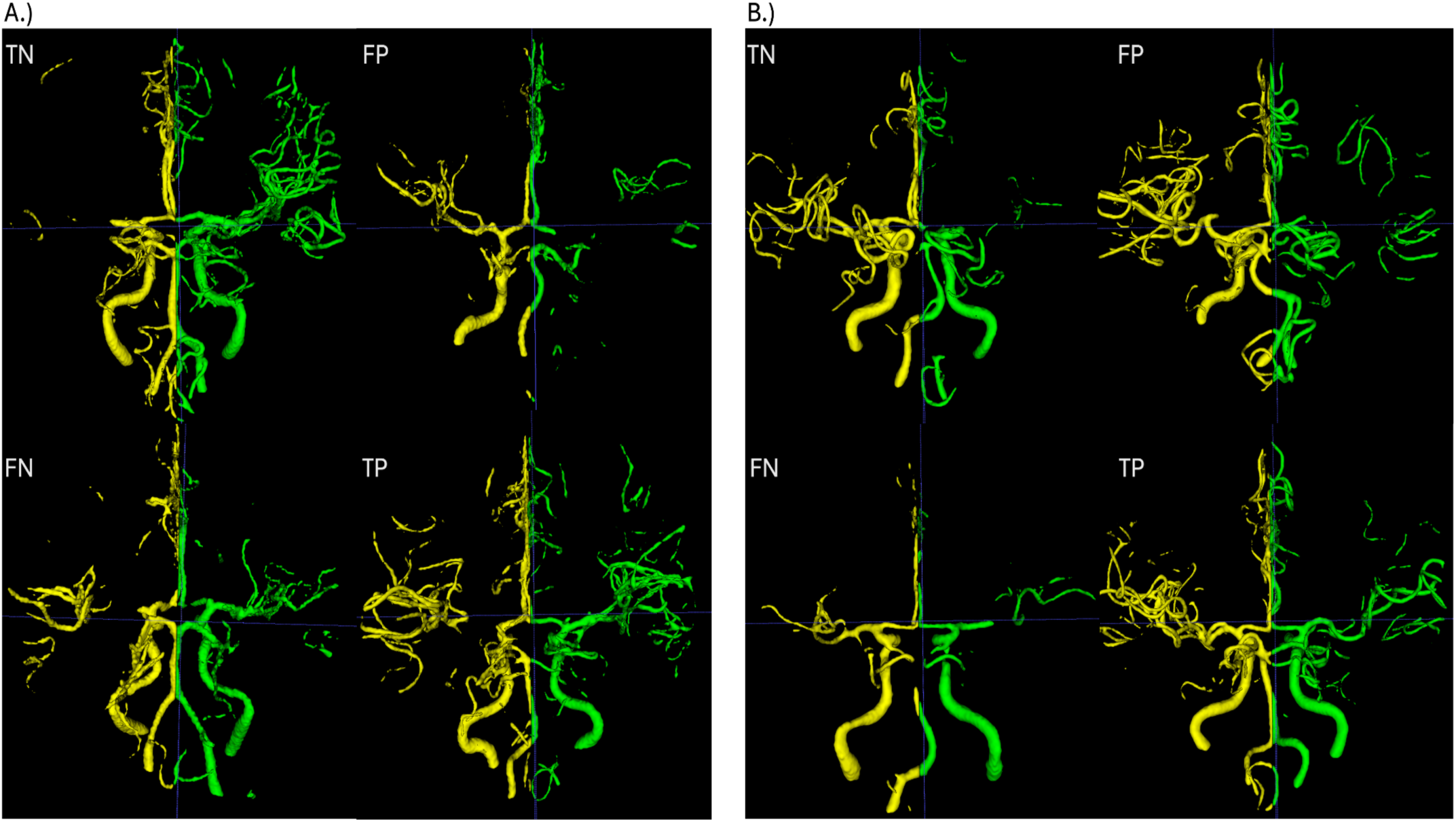
Error analysis of Model C (Vessels + CoW) result examples: A.) Results from the internal test set. B.) Results from the external test set.

#### MCA masks ROI

For the MCA mask model B, the radiomics selection pipeline identified a total of 97 predictive features. In the internal test set, the model demonstrated an average AUC of 0.8157 (95% CI: [0.707– 0.909]; True Positives: 45, True Negatives: 10, False Positives: 13, False Negatives: 1). In the external test set, the model achieved an AUC of 0.663 (95% CI: [0.554–0.765]; True Positives: 74, True Negatives: 20, False Positives: 21, False Negatives: 25).

#### Combination of binary and CoW vessel segmentation ROIs

Although the model C performance on the internal test set was slightly lower compared to the model using only the shape features from the vessel trees (Test AUC: 0.8611, 95% CI: 0.7484–0.9490), it performed better on the external test set, achieving a Test AUC of 0.8677 (95% CI: 0.7983–0.9251). This made it the best-performing model on the external test set. *Performance metrics are summarized in Table 3*.

**Table 3:**
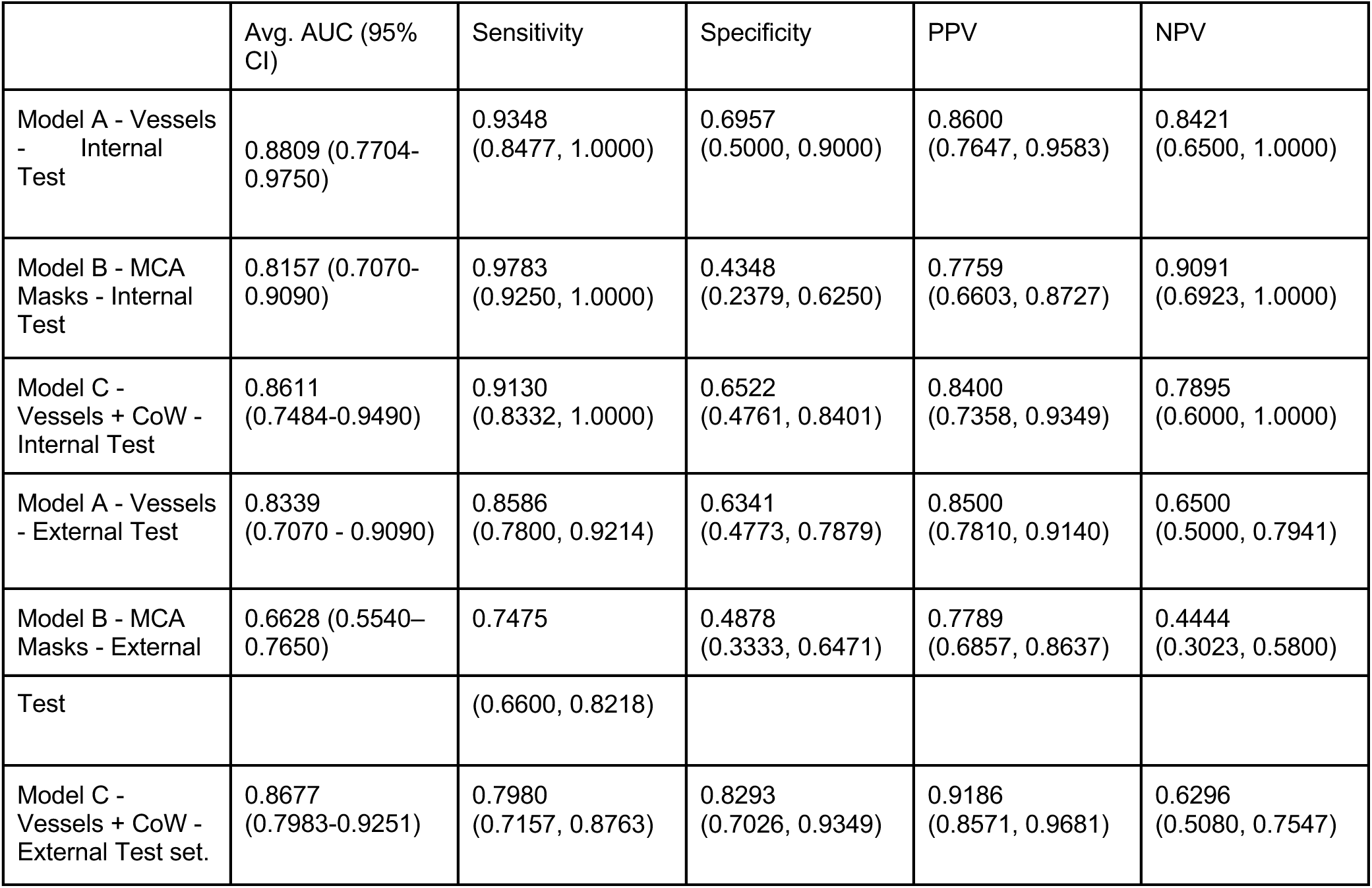
Comparison of AUC performance between models using vessel masks and MCA masks on the internal and external test sets. On the internal test set, the difference was statistically significant (DeLong’s test, p = 0.003), and this was confirmed on the external test set (p < 0.001). Additionally, comparison between the model using vessel features alone and the model combining vessel and Circle of Willis (CoW) features showed significant improvement on both the internal (p = 0.0312) and external (p = 0.0062) test sets (DeLong’s test for AUC)

|  | Avg. AUC (95% CI) | Sensitivity | Specificity | PPV | NPV |
| --- | --- | --- | --- | --- | --- |
| Model A - Vessels - Internal Test | 0.8809 (0.7704-0.9750) | 0.9348 (0.8477, 1.0000) | 0.6957 (0.5000, 0.9000) | 0.8600 (0.7647, 0.9583) | 0.8421 (0.6500, 1.0000) |
| Model B - MCA Masks - Internal Test | 0.8157 (0.7070-0.9090) | 0.9783 (0.9250, 1.0000) | 0.4348 (0.2379, 0.6250) | 0.7759 (0.6603, 0.8727) | 0.9091 (0.6923, 1.0000) |
| Model C - Vessels + CoW - Internal Test | 0.8611 (0.7484-0.9490) | 0.9130 (0.8332, 1.0000) | 0.6522 (0.4761, 0.8401) | 0.8400 (0.7358, 0.9349) | 0.7895 (0.6000, 1.0000) |
| Model A - Vessels - External Test | 0.8339 (0.7070 - 0.9090) | 0.8586 (0.7800, 0.9214) | 0.6341 (0.4773, 0.7879) | 0.8500 (0.7810, 0.9140) | 0.6500 (0.5000, 0.7941) |
| Model B - MCA Masks - External | 0.6628 (0.5540-0.7650) | 0.7475 | 0.4878 (0.3333, 0.6471) | 0.7789 (0.6857, 0.8637) | 0.4444 (0.3023, 0.5800) |
| Test |  | (0.6600, 0.8218) |  |  |  |
| Model C -<br>Vessels + CoW -<br>External Test set. | 0.8677<br>(0.7983-0.9251) | 0.7980<br>(0.7157, 0.8763) | 0.8293<br>(0.7026, 0.9349) | 0.9186<br>(0.8571, 0.9681) | 0.6296<br>(0.5080, 0.7547) |

## Discussion

Our study presents a novel cerebrovascular radiomics approach designed to automate, objectify and improve the assessment of collateral status in patients with acute ischemic stroke. To the best of our knowledge, this is the first approach to leverage radiomic features extracted from vessel segmentations for collateral scoring on CTA.

Unlike recent studies that rely on advanced imaging such as multiphase CTA ^32^ or MR perfusion ^33^ our pipeline utilises CTA, a modality widely used across stroke centers, ensuring both practicality and scalability in diverse clinical settings.

In contrast to other radiomics studies on CTA, we evaluated our approach using multi-center internal and external datasets. Results demonstrate that cerebrovascular radiomics substantially outperform those limited to the established MCA atlas-based methods ^18^. This is likely due to the broader vascular context captured by vessel segmentation, which includes contributions from the anterior and posterior circulations. These territories are essential for collateral recruitment, as better perfusion in these regions may reflect more robust leptomeningeal anastomoses and compensatory pathways.

Additionally, we hypothesize that the improved performance achieved by incorporating radiomic features from the CoW can be attributed to capturing anatomical variations in the communicating arteries. This hypothesis is based on the fact that the most predictive radiomic feature (VoxelVolume) for the task was derived from the region corresponding to the communicating segments of the Circle of Willis, This finding is consistent with prior studies highlighting the clinical relevance of CoW configuration, particularly in relation to thrombus location and collateral recruitment ^34,35^.

In the acute clinical setting, conventional collateral scoring relies on manual interpretation by neuroradiologists, a process subject to interrater variability with only moderate agreement among experts (κ-values between 0.45 and 0.65) ^36^. Our pipeline can be easily integrated in the clinical routine and resolve discrepancies between neuroradiologists and therefore assist collateral status-based intervention decision-making ^5^.

In addition to the acute clinical routine, our work’s utility extends to objectively improving retrospective datasets. Many stroke outcome prediction models have been trained on retrospective data, with collateral status often being included as a critical factor ^37^. However, these models are typically derived from multiple centers worldwide, where inter-rater variability in collateral assessment is common. This variability can diminish the predictive value of these models. Therefore, by standardizing collateral status evaluation using our systematized method, the accuracy and performance of existing predictive models may be enhanced, ensuring more reliable models in stroke outcome research ^38^.

Beyond collateral status assessment, our vessel segmentation pipeline lays the foundation for broader applications in neurovascular care. For instance, accurate vascular quantification could improve the detection of large vessel occlusions, a known challenge for current deep-learning models in real-world settings ^39^. Furthermore, detailed morphological analysis of crucial vessel characteristics such as diameter ^40^ and tortuosity ^41^ could guide the selection of catheters, stent retrievers, and aspiration devices and tailor EVT strategies to optimize recanalization success.

### Limitations and Prediction Error analysis

This study reveals several noteworthy limitations concerning the dataset. First, it exclusively focuses on proximal anterior circulation large vessel occlusions (LVOs). Consequently, the models have not been exposed to the full clinical spectrum of LVO presentations, including distal and posterior circulation occlusions, which diminishes the overall generalizability of the findings.

Additionally, a significant variability in the timing of the contrast bolus across the CTA acquisitions was observed. In instances where imaging occurred during the early arterial or late venous phase, the status of the arterial vessel may have been misinterpreted, potentially leading to misleading predictions regarding collateral status (please refer to (Figure 7: B for an example of a false negative). Another crucial limitation lies in the manual assessment of collateral status. We employed the Tan collateral score, which ranges from 0 to 3, as our reference standard. However, distinguishing between intermediate grades—especially grade 1 and grade 2—often proves subjective, as quantifying the filing status accurately as a relative percentage (>50% or <50%) can be challenging (see Figure 7 B for a false positive example and Figure 7 A for a false negative example).

Moreover, the quality of the segmentations plays a vital role in the models’ overall performance. While minor inaccuracies were noted, visual inspections confirmed the absence of significant errors. A further limitation of our approach pertains to the division of the vascular tree into left and right hemispheres. This method, lacking a side-specific anatomical model, can be influenced by individual variations in vascular anatomy. Consequently, vessels that anatomically belong to one hemisphere may inadvertently be misassigned to the other, which can negatively impact both feature extraction and predictive accuracy (see Figure 7 A for an example of a false positive). In light of these considerations, it is essential to devote more effort to the development of robust anatomical vessel segmentation models and to thoroughly evaluate their performance.

## Conclusion

In summary, our study shows the potential of vessel-specific segmentation combined with radiomics analysis to quantify collateral status. By focusing on precise vascular structures rather than broad anatomical regions and by automatizing the collateral scoring process, our approach offers a more precise and generalizable alternative to current methodologies. While few limitations remain, our work lays the groundwork for collateral status assessment by integrating vessel analysis into routine clinical workflows, ultimately aiming to personalize and optimize stroke management strategies.

## Data Availability

The imaging and clinical data from the MR CLEAN trial and the external dataset provided by University Hospital Heidelberg are not publicly available due to privacy and data-sharing restrictions. However, the complete code for data preprocessing, vessel segmentation, radiomics feature extraction, feature selection, and classification is available open-source in a public GitHub repository:https://github.com/claim-berlin/CollateralScore

## Acknowledgements

None.

## Abbreviations

NIfTI: Neuroimaging Informatics Technology Initiative
CTA: Computer Tomography Angiography
LVO: Large Vessel Occlusion
DICOM: Digital Imaging and communication in Imaging
mRS: modified Rankin Scale
EVT: Endovascular thrombectomy
AUC: Area Under the Curve
HU: Hounsfield’s Units
ROI: Region of Interest
NIHSS: National Institutes of Health Stroke Scale
ICA: Internal Carotid Artery
MCA: Middle Cerebral Artery
ACA: Anterior Cerebral Artery
AcoA: Anterior Communicating Artery
PcoA: Posterior Communicating Artery
PCA: Posterior Cerebral Artery
AUC: Area Under the Receiver Operating Characteristic Curve

## References

1. Malhotra, K., Gornbein, J. & Saver, J. L. Ischemic Strokes Due to Large-Vessel Occlusions Contribute Disproportionately to Stroke-Related Dependence and Death: A Review. Front. Neurol. 8, 651 (2017).

2. Sanelli, P. C. et al. Imaging and Treatment of Patients with Acute Stroke: An Evidence-Based Review. *Am*. J. Neuroradiol. 35, 1045–1051 (2014).

3. Mangiardi, M. et al. The Pathophysiology of Collateral Circulation in Acute Ischemic Stroke. Diagnostics 13, 2425 (2023).

4. Maguida, G. & Shuaib, A. Collateral Circulation in Ischemic Stroke: An Updated Review. J. Stroke 25, 179–198 (2023).

5. Huijberts, I. et al. Collateral-based selection for endovascular treatment of acute ischaemic stroke in the late window (MR CLEAN-LATE): 2-year follow-up of a phase 3, multicentre, open-label, randomised controlled trial in the Netherlands. Lancet Neurol. 23, 893–900 (2024).

6. Goyal, M. et al. Endovascular Treatment of Stroke Due to Medium-Vessel Occlusion. N. Engl. J. Med. 392, 1385–1395 (2025).

7. Nambiar, V. et al. CTA Collateral Status and Response to Recanalization in Patients with Acute Ischemic Stroke. AJNR Am. J. Neuroradiol. 35, 884–890 (2014).

8. Berkhemer, O. A. et al. A Randomized Trial of Intraarterial Treatment for Acute Ischemic Stroke. N. Engl. J. Med. 372, 11–20 (2015).

9. Boers, A. M. M. et al. Value of Quantitative Collateral Scoring on CT Angiography in Patients with Acute Ischemic Stroke. AJNR Am. J. Neuroradiol. 39, 1074–1082 (2018).

10. Mair, G. et al. Observer reliability of CT angiography in the assessment of acute ischaemic stroke: data from the Third International Stroke Trial. Neuroradiology 57, 1–9 (2015).

11. Fasen, B. A. C. M., Heijboer, R. J. J., Hulsmans, F.-J. H. & Kwee, R. M. CT Angiography in Evaluating Large-Vessel Occlusion in Acute Anterior Circulation Ischemic Stroke: Factors Associated with Diagnostic Error in Clinical Practice. AJNR Am. J. Neuroradiol. 41, 607–611 (2020).

12. Sichtermann, T. et al. Deep Learning–Based Detection of Intracranial Aneurysms in 3D TOF-MRA. *Am*. J. Neuroradiol. 40, 25–32 (2019).

13. Hilbert, A. et al. BRAVE-NET: Fully Automated Arterial Brain Vessel Segmentation in Patients With Cerebrovascular Disease. *Front*. Artif. Intell. 3, (2020).

14. Dong, M. et al. Artificial intelligence-based automatic nidus segmentation of cerebral arteriovenous malformation on time-of-flight magnetic resonance angiography. Eur. J. Radiol. 178, 111572 (2024).

15. van Griethuysen, J. J. M. et al. Computational Radiomics System to Decode the Radiographic Phenotype. Cancer Res. 77, e104–e107 (2017).

16. A quantitative symmetry-based analysis of hyperacute ischemic stroke lesions in noncontrast computed tomography - PMC. https://pmc.ncbi.nlm.nih.gov/articles/PMC5339891/.

17. Ramos, L. A. et al. Combination of Radiological and Clinical Baseline Data for Outcome Prediction of Patients With an Acute Ischemic Stroke. Front. Neurol. 13, (2022).

18. Avery, E. W. et al. Radiomics-Based Prediction of Collateral Status from CT Angiography of Patients Following a Large Vessel Occlusion Stroke. Diagnostics 14, 485 (2024).

19. Hoopes, A., Mora, J. S., Dalca, A. V., Fischl, B. & Hoffmann, M. SynthStrip: skull-stripping for any brain image. NeuroImage 260, 119474 (2022).

20. Muschelli, J. A Publicly Available, High Resolution, Unbiased CT Brain Template. in Information Processing and Management of Uncertainty in Knowledge-Based Systems (eds Lesot, M.-J. et al.) 358–366 (Springer International Publishing, Cham, 2020). doi:10.1007/978-3-030-50153-2_27.

21. Isensee, F., Jaeger, P. F., Kohl, S. A. A., Petersen, J. & Maier-Hein, K. H. nnU-Net: a self-configuring method for deep learning-based biomedical image segmentation. Nat. Methods 18, 203–211 (2021).

22. Tan, I. Y. L. et al. CT Angiography Clot Burden Score and Collateral Score: Correlation with Clinical and Radiologic Outcomes in Acute Middle Cerebral Artery Infarct. AJNR Am. J. Neuroradiol. 30, 525–531 (2009).

23. Srivatsa, S. et al. Cerebral vessel anatomy as a predictor of first-pass effect in mechanical thrombectomy for emergent large-vessel occlusion. J. Neurosurg. 134, 576–584 (2020).

24. van Griethuysen, J. J. M. et al. Computational Radiomics System to Decode the Radiographic Phenotype. Cancer Res. 77, e104–e107 (2017).

25. Tejani, A. S. et al. Checklist for Artificial Intelligence in Medical Imaging (CLAIM): 2024 Update. *Radiol*. Artif. Intell. 6, e240300 (2024).

26. Kocak, B. et al. CheckList for EvaluAtion of Radiomics research (CLEAR): a step-by-step reporting guideline for authors and reviewers endorsed by ESR and EuSoMII. Insights Imaging 14, 75 (2023).

27. Li, K., Wang, F., Yang, L. & Liu, R. Deep Feature Screening: Feature Selection for Ultra High-Dimensional Data via Deep Neural Networks. Preprint at 10.48550/arXiv.2204.01682 (2023).

28. Pedregosa, F. et al. Scikit-learn: Machine Learning in Python. Mach. Learn. PYTHON.

29. Breiman, L. Random Forests. Mach. Learn. 45, 5–32 (2001).

30. Akiba, T., Sano, S., Yanase, T., Ohta, T. & Koyama, M. Optuna: A Next-generation Hyperparameter Optimization Framework. in Proceedings of the 25th ACM SIGKDD International Conference on Knowledge Discovery & Data Mining 2623–2631 (Association for Computing Machinery, New York, NY, USA, 2019). doi:10.1145/3292500.3330701.

31. Chen, W. et al. On the assessment of the added value of new predictive biomarkers. BMC Med. Res. Methodol. 13, 98 (2013).

32. Liu, H. et al. Deep Learning based Collateral Scoring on Multi-Phase CTA in patients with acute ischemic stroke in MCA region. *Am*. J. Neuroradiol. (2025) doi:10.3174/ajnr.A8911.

33. Tetteh, G. et al. A deep learning approach to predict collateral flow in stroke patients using radiomic features from perfusion images. Front. Neurol. 14, 1039693 (2023).

34. Thirugnanachandran, T. et al. Anterior Cerebral Artery Stroke: Role of Collateral Systems on Infarct Topography. Stroke 52, 2930–2938 (2021).

35. Casault, C. et al. Collateral Scoring on CT Angiogram Must Evaluate Phase and Regional Pattern. Can. J. Neurol. Sci. 44, 503–507 (2017).

36. Wolff, L. et al. Inter-rater reliability for assessing intracranial collaterals in patients with acute ischemic stroke: comparing 29 raters and an artificial intelligence-based software. Neuroradiology 64, 2277–2284 (2022).

37. Venema, E. et al. Prediction of Outcome and Endovascular Treatment Benefit: Validation and Update of the MR PREDICTS Decision Tool. Stroke 52, 2764–2772 (2021).

38. Liu, Y. et al. Functional Outcome Prediction in Acute Ischemic Stroke Using a Fused Imaging and Clinical Deep Learning Model. Stroke 54, 2316–2327 (2023).

39. Real world clinical experience of using Brainomix e-CTA software in a medium size acute National Health Service Trust | British Journal of Radiology | Oxford Academic. https://academic.oup.com/bjr/article/98/1168/592/7989295.

40. Tanaka, Y. et al. Optimizing thrombectomy in medium vessel occlusion: Focus on vessel diameter. Interv. Neuroradiol. J. Peritherapeutic Neuroradiol. Surg. Proced. Relat. Neurosci. 15910199241272638 (2024) doi:10.1177/15910199241272638.

41. Bala, F. et al. Impact of vessel tortuosity and radiological thrombus characteristics on the choice of first-line thrombectomy strategy: Results from the ESCAPE-NA1 trial. Eur. Stroke J. 8, 675–683 (2023).

